# A Comparative Analysis of Electronic Health Record and Electrocardiogram Waveform Data for Pulmonary Embolism Identification in Critically Ill Patients

**DOI:** 10.1101/2025.09.24.25335530

**Authors:** Sampath Rapuri, Carl Harris, Kirby Gong, Robert D. Stevens

**Author notes:** Corresponding Author: Robert D. Stevens (RS).

## Abstract

Pulmonary embolism (PE) is one of the leading causes of preventable death amongst hospitalized patients, yet current risk assessment tests based on clinical variables have shown inconsistent validity or poor predictiveness. More recent predictive models for PE using electronic health record (EHR) data are promising, but their reliance on comprehensive and integrated EHR data can limit real-time utility, creating a need for more accessible and rapid diagnostic tools. This study compares the performance of an EHR-based model, an electrocardiogram waveform (WF) model, and a fusion model combining both modalities for the identification of PE in critically ill patients. We leverage routinely acquired clinical and ECG waveform data from the 48 hours preceding PE suspicion from a retrospective dataset of 7,132 ICU admissions between 2008 and 2019 (4.60% PE prevalence). PE diagnoses were determined through ICD-9 or ICD-10 diagnostic coding. We find that our WF model, which employs a single, 10-second 12-lead ECG sample, demonstrated comparable predictive performance (AUROC 0.67 (95% CI, 0.64–0.70)) to our EHR-based model (AUROC 0.71 (0.68–0.74)). However, a fusion model combining both modalities did not improve predictive performance (AUROC 0.67 (0.64–0.70)). All our models outperform widely used existing risk stratification scores such as the Revised Geneva score (AUROC 0.54 (0.51–0.57)), the original Wells score (AUROC 0.61 (0.58–0.64)), and the PE Rule Out Criteria (AUROC 0.56 (0.53–0.59)). Our findings underscore the value that ECG waveform data can bring to the detection of PE in critically ill patients by demonstrating its predictive capability compared to existing benchmarks. After additional validation, these models may serve as valuable tools in PE diagnostic clinical workflows.

**Author Summary:** Pulmonary embolism (PE) is a life-threatening condition resulting from an embolus that obstructs blood flow in the arteries of the lung. Although recent advancements in the treatment of PE have improved patient outcomes and reduced mortality, existing risk scoring systems still lack discriminatory power and fail to validate in specialized populations like those in the Intensive Care Unit (ICU). In this cohort study, we developed PE detection models for critically ill patients using a large, open-source clinical dataset that outperforms current benchmark risk stratification scores. Our approach leverages ECG waveform data, obtained at least 48 hours before clinical suspicion of PE, potentially enabling earlier therapeutic intervention. Furthermore, by incorporating hand-crafted features during model training, our study provides detailed insights into some predictive factors of PE derived from ECG waveform data.

## Introduction

Pulmonary embolisms are obstructions of blood flow to the lungs, belonging to a wider class of venous disorders called venous thromboembolisms (VTEs). These blockages typically result from emboli originating from a deep vein thrombosis (DVT) in the lower extremities, though they can arise from other parts of the body (1). Collectively, DVTs and PEs are life-threatening conditions with a high risk of recurrence and chronic complications. In the United States alone, approximately 900,000 individuals are diagnosed annually, and up to 30% of these patients die within one month of diagnosis (2–4). The high mortality rate is further compounded in the difficulty of diagnosis as PEs often mimic other common illnesses, leading to frequent misdiagnoses in up to 50% of patients (5). Consequently, early detection and treatment are critical, as with prompt intervention, patient mortality rates can be reduced from 30% to 8% (6). These diagnostic challenges highlight the urgent need for more accurate and reliable tools for risk assessment and prediction.

Because of the difficulty diagnosing PE, physicians are often forced to rely on resource intensive or specialized tests such as D-dimer measurements or computed tomography pulmonary angiogram (CTPA), potentially resulting in unnecessary resource utilization, patient harm, and increased medical expenses (7). To aid in the diagnostic workflow, several risk stratification scores have been proposed such as the Wells Scoring System (8), Revised Geneva Scoring System (9), and the Pulmonary Embolism Rule Out Criteria (10). However, the performance of these models is not consistent across different patient populations and demonstrates poor discrimination and specificity in inpatient populations where it is essential due to the heightened vulnerability and complexity of these patients’ conditions (11,12). Previous studies have developed improved PE prediction models leveraging electronic health record (EHR) data that outperform common risk stratification scores in a broad population of hospitalized and ICU patients and demonstrate strong generalizability when validated with external hospital datasets from different populations (13,14).

While these EHR-based models are promising, their reliance on comprehensive, aggregated data can limit their utility for rapid, point-of-care risk stratification. The electrocardiogram (ECG) presents a compelling alternative, as it is a low-cost, rapid, and widely available diagnostic modality in critical care settings. Several ECG abnormalities, including right ventricular strain patterns, the S1Q3T3 pattern, and T-wave inversions, have previously been reported as useful indicators for predicting mortality in PE patients (15–17). Additional research has further demonstrated that ECG waveforms can be used to predict PE but these prior studies faced limited actionability as they utilized narrow predictive windows or were not as performant without integration with EHR data (18–20). We hypothesize that a model relying solely on a 12-lead ECG can match the predictive performance of a model using comprehensive EHR data. We also test whether a fusion model that integrates both data sources can outperform the individual unimodal models.

In this study, we train models to identify PE in critically ill patients utilizing widely available 500 Hz, 10 second, 12-lead ECG waveforms and patient medical records. We compare WF, EHR-based, and fusion models to identify the advantages of each approach and determine whether combining both modalities improves predictive performance. We further leverage a large, open-source dataset to ensure broad applicability and transparency of this work.

## Methods

### Dataset and study population

We developed and evaluated our models using the MIMIC-IV (v2.2) dataset, which is a large open-source dataset that includes 299,712 patients across 431,231 hospital admissions to the Beth Israel Deaconess Medical Center between 2008 and 2019 (21).

We identify our PE cohort using ICD 9/10 diagnostic codes (listed in Supplementary table S1). However, the MIMIC-IV dataset does not record the time of diagnosis – this means that we cannot definitively determine at what point in time during a patient’s hospital admission the PE was identified or recorded. Instead, we rely on the time of the first recorded chest CT scan during a patient’s hospital admission as a surrogate marker for the onset of clinical suspicion, using data from 48 hours and earlier from the time of suspicion for our risk stratification model. Records of D-dimer test administration were also considered; however, given the low prevalence of these recordings in our cohort, using the D-dimer test time was not adopted. It is important to explicitly state that because of this methodology, our model is developed and validated on a moderate- to high suspicion PE cohort, which may not generalize as a screening tool in the broader ICU population

Using these criteria, we identified 335 ICU admissions with a recorded diagnosis of pulmonary embolism (4.63% PE prevalence) and 6,906 ICU admissions without such a diagnosis. Because ECG data quality can be variable, we excluded patients with any missing lead II data, which accounted for less than 2% of the overall cohort. In addition, we limited our analysis to an adult population. The complete inclusion and exclusion criteria are detailed in Figure 1.

**Figure 1.**
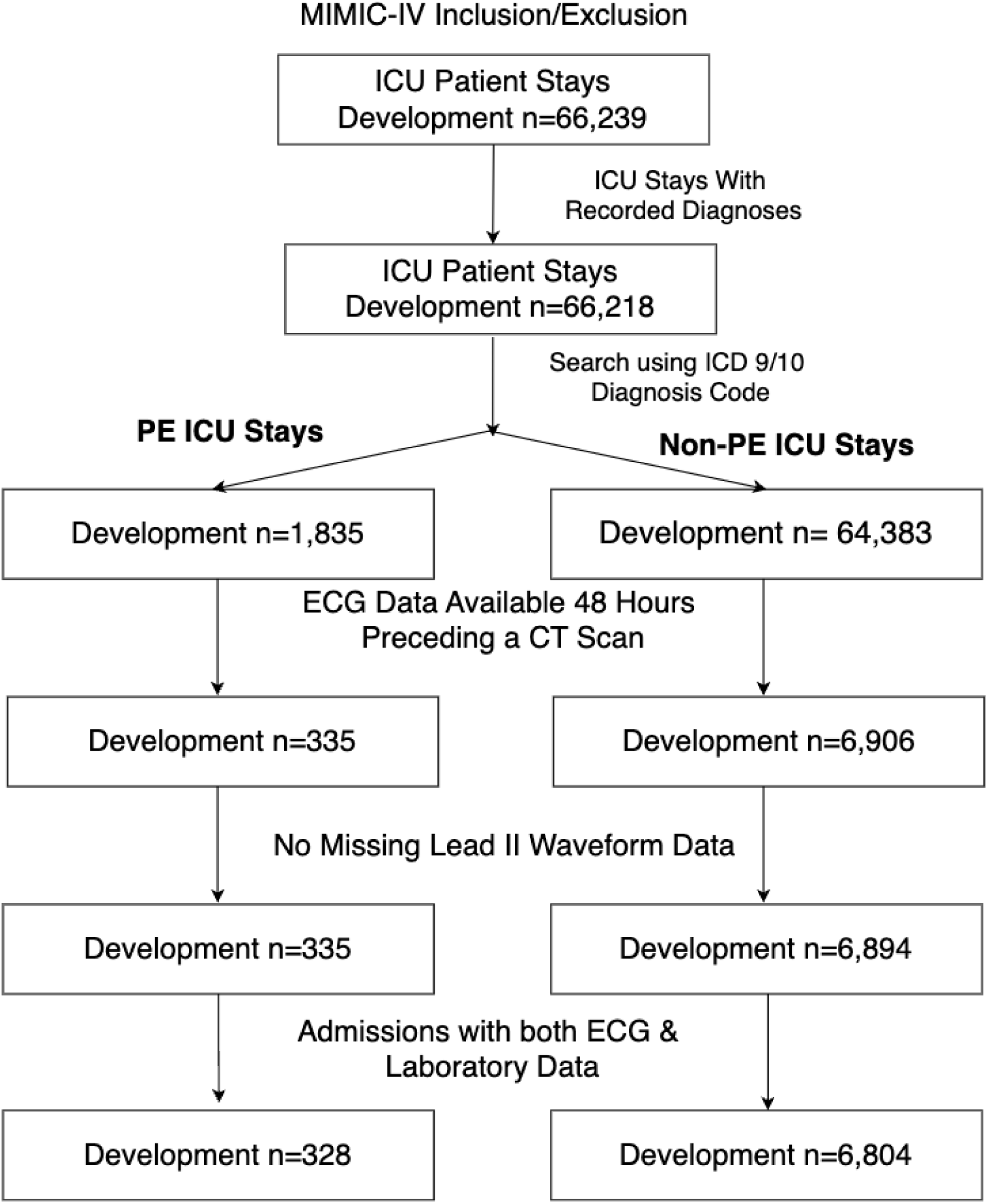
Inclusion & Exclusion criteria employed in this study.

To ensure replicability and transparency of this work, we follow the Strengthening the Reporting of Observational Studies in Epidemiology (STROBE) and the Transparent Reporting of a Multivariable Prediction Model for Individual Prognosis or Diagnosis (TRIPOD) checklists (22,23) (Supplementary tables S2 and S3).

### Predictive Variables & Feature Extraction

Model variables were selected based on a review of the literature and clinician input. Our objective was to develop a model for early risk stratification in critically ill patients regardless of initial PE suspicion. To this end, we limited our model inputs to clinical variables routinely collected in at least 75% of our selected cohort. Our extracted EHR variables include patient demographics, medical history, laboratory studies, comorbidities, low-frequency physiologic time series (e.g., heart rate and blood pressure), medications administered, and fluid input/output records. Comorbidities are extracted using the R package ‘comorbidity’ (24). From numerical time series data, we extract estimates of central tendency (mean, median, and standard deviation), regression-related features (slopes and intercepts), minimum, maximum, first, and last values. Non-time series numerical data were not processed further. Lastly, categorical variables were one-hot encoded.

Waveform features included ECG morphological features, time-domain, frequency-domain, nonlinear heart-rate variability features, general signal processing features, ECG-derived respiration rate variability features, and vectorcardiogram features. Waveform feature extraction was performed using the WFDB (25), Neurokit2 (26), and Tsfresh (27) packages. Vectorcardiogram feature extraction was performed using the BRAVEHEART (28) package in Matlab R2024a.

All exploratory data analysis was conducted using the NumPy (29) and Pandas (30) packages in Python 3.11. A full list of waveform and non-waveform features explored in this study are also available in the supplementary material (Supplementary tables S3 and S4).

### Preprocessing

Non-waveform numerical feature distributions were manually adjusted via input from a fellowship-trained, board-certified intensive care physician who defined upper and lower bounds of physiologic plausibility. Values outside of these set ranges were imputed using training data median values. One-hot encoded features were imputed using the most commonly occurring values from the training data. We preprocess our ECG waveform data using a bandpass filtering strategy described in Pan and Tompkins (31). For our waveform features, missing non-lead II waveform data are linearly interpolated, while ECG morphological features from the MIMIC-IV dataset are imputed using training data median values. All features are finally normalized and scaled using the Scikit-learn package (32).

### Modeling

We developed three models to detect PE: a WF model trained on ECG waveform features, an EHR model trained on routinely collected data such as vital signs, laboratory studies, and demographics, and a fusion model that integrates both data modalities. We evaluated several modeling approaches for all models, including logistic regression, random forest, and ensemble methods such as XGBoost (33), and CatBoost (34). For our fusion model, we leveraged the predicted PE probabilities from the base EHR and WF classifiers as input features within a stacked classifier framework.

Given the relatively low number of PE patients, we implemented a rigorous feature selection process to mitigate potential concerns of overfitting. For EHR data, we first used a recursive feature elimination (RFE) approach with an XGBoost model (using default hyperparameters) to remove features with zero importance and then applied mutual information (MI)–based feature selection to select features that provided the most information about the target label. In contrast, when we applied RFE-based feature selection to the waveform-derived features, we observed a significant drop in model performance, as measured by validation AUROCs. This suggests that RFE might be discarding features that, although not highly ranked individually, collectively contribute valuable predictive information. Therefore, for the waveform feature space, we relied solely on MI-based feature selection to preserve these informative features.

A nested cross-validation scheme was employed to evaluate model performance and tune hyperparameters using a Bayesian search implemented with the Optuna package (35). Initially, the data was divided into five outer folds, with each fold stratified by PE target labels to ensure a consistent representation of PE patients. For every outer fold, the training portion was further partitioned into three inner splits—again stratified by PE target labels—with two splits designated for training and one reserved for validation. The AUROC served as the primary metric for assessing model performance and selecting the optimal hyperparameters for each inner fold. This nested approach is critical as it ensures that hyperparameter tuning is performed on data separate from the final evaluation on the withheld test set, providing an unbiased estimate of model generalizability.

For each model, 500 hyperparameter combinations were evaluated within each outer fold, resulting in a total of 2,500 unique hyperparameter combinations across all outer folds. After hyperparameter tuning, each model was trained on the entire training set of the outer fold with the best-selected hyperparameters and evaluated on the respective held-out test set. We aggregated the AUROC scores across all outer folds to assess overall performance and variability. The uncertainty in our performance estimates is estimated by bootstrapping with 10,000 resamples to obtain 95% confidence intervals.

### Model Performance Evaluation

We report performance using three non-thresholded metrics—AUROC, area under the precision-recall curve (AUPRC), and the Brier score—and three threshold-dependent metrics, including negative predictive value (NPV), positive predictive value (PPV), and specificity at a fixed sensitivity of 0.80. All performance metrics for the three models developed in this study, evaluated on the testing data, are presented with their corresponding 95% confidence intervals. The AUROC reflects the model’s predictiveness by describing the trade-off between the true positive and false positive rates across various thresholds. In contrast, the AUPRC focuses on the balance between precision and recall, making it particularly informative for datasets with class imbalances such as ours. The NPV and PPV can measure the reliability of a model’s predictions for negative and positive outcomes, respectively, while the Brier score quantifies the model calibration. χ^2^ tests of independence and Mann-Whitney U tests are used to compare differences between groups. During statistical testing, we conducted a permutation test with 10,000 iterations and then adjusted the significance threshold using a Bonferroni correction (36) to account for the multiple comparisons. Statistical analyses were performed using the Scipy package (37).

### Feature Analysis

Feature importances were determined by calculating SHapley Additive exPlanations (Shapley values) (38). Shapley values offer a robust method for quantifying the relative contribution of each feature to the model’s output. During our cross-validation procedure, we computed the mean absolute SHAP values for each of the three models on the outer testing folds and then aggregated these values across all folds. This approach yielded consolidated estimates that reflect both the average feature contributions and their variability across different data splits.

### Patient characteristics

A total of 66,239 ICU admissions were analyzed in this study, with 7,241 meeting our inclusion criteria of having recorded ECGs within the 48 hours preceding a chest CT scan, as illustrated in Figure 1. After excluding admissions with missing lead II data, we obtained a cohort of 7,132 ICU admissions, of which 4.60% were diagnosed with a PE. ICU admissions with PE had a higher inter-hospital transfer rate (25.30% vs. 19.55%) and a lower rate of Emergency Department admissions (58.54% vs. 65.71%) compared with non-PE ICU admissions. Comorbid conditions such as fluid and electrolyte disorders (37.20%), chronic pulmonary disease (21.95%), and coagulopathy (14.33%) were significantly more prevalent in the PE group, highlighting the complex clinical profiles of critically ill PE patients. Furthermore, PE ICU admissions experienced a longer median length of stay (117.84 hours) and slightly higher in-ICU mortality rates (19.51% vs. 18.56%) compared with those without PE. A summary and comparison of patient characteristics between the PE and non-PE groups is provided in Table 1.

**Table 1.**
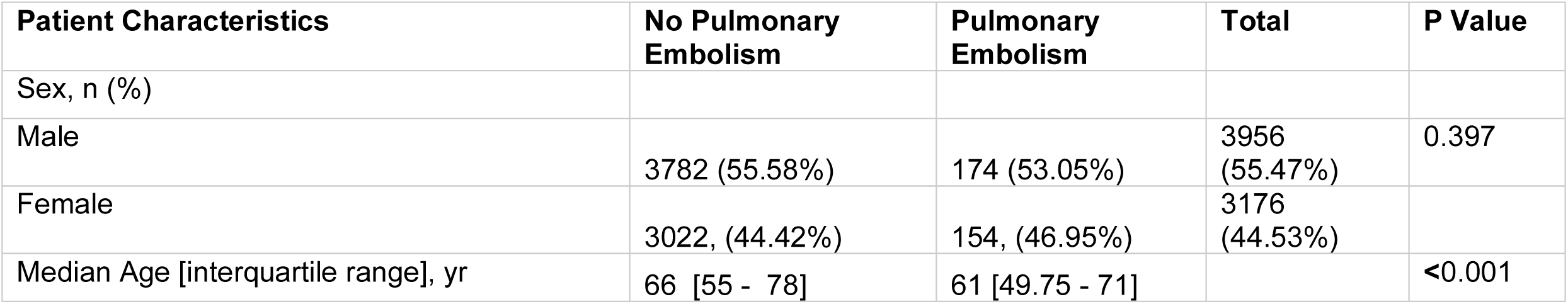
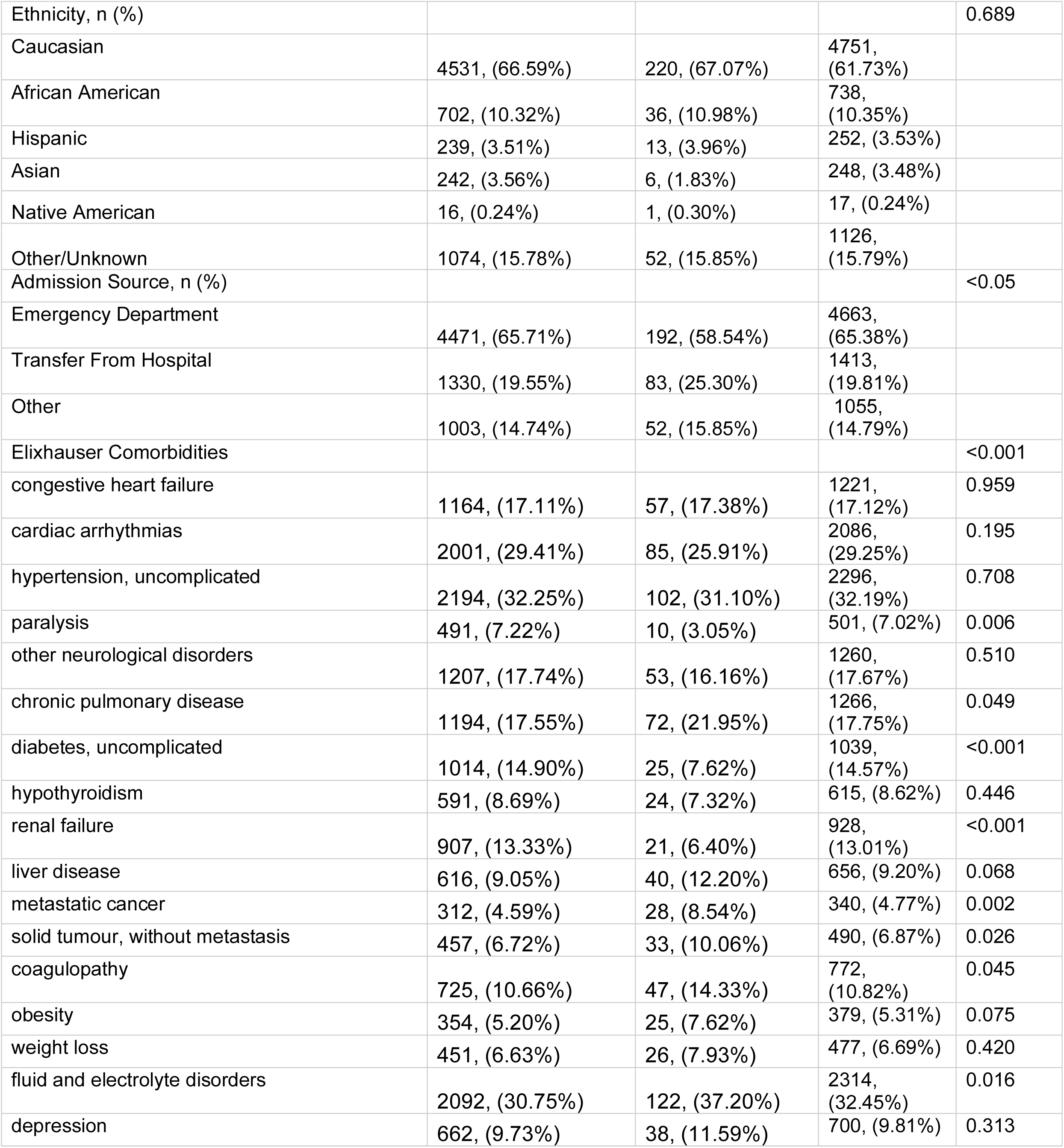
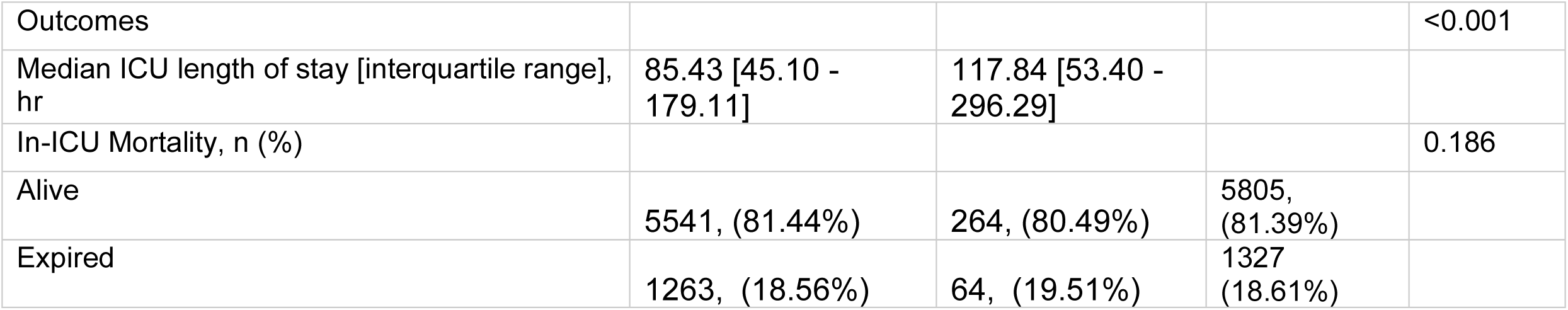
Patient Characteristics in the development dataset.

### Model Performance Evaluation

Model performances for the development dataset are presented in Table 2. The EHR model achieved an AUROC of 0.71 (0.68–0.74), while the WF model achieved a comparable AUROC of 0.67 (0.64–0.70). The fusion model performs comparably to both models, achieving an AUROC of 0.71 (0.68–0.74). Likewise, the AUPRC of all three models is similar, with the fusion model attaining the highest AUPRC of 0.13 (0.11–0.16). Figures 2. and 3. display the AUROC and AUPRC for all models in this study, respectively. The EHR and fusion models significantly (*p* < 0.001) outperform baseline models based on waveform architectures proposed by Wysokinski et al. (19)., Somani et al. (20), and Silva et al. (39), which were trained using the same training and evaluation pipelines as our model. Similarly, the EHR and fusion models outperform all baseline EHR models (*p* < 0.001). Our WF model outperforms Silva et al. (39), the Pulmonary Embolism Rule Out Criteria (10), and the Revised Geneva score (9) (*p* < 0.001). The WF model outperforms Somani et al. (20) and the Original Wells score (8) but not significantly (*p*=0.009 and *p=*0.007, respectively). Supplementary tables S5, S6, and S7 detail the performance of the WF, EHR, and fusion models across all evaluated models (logistic regression, random forest, XGBoost (33), and CatBoost (34)).

**Figure 2.**
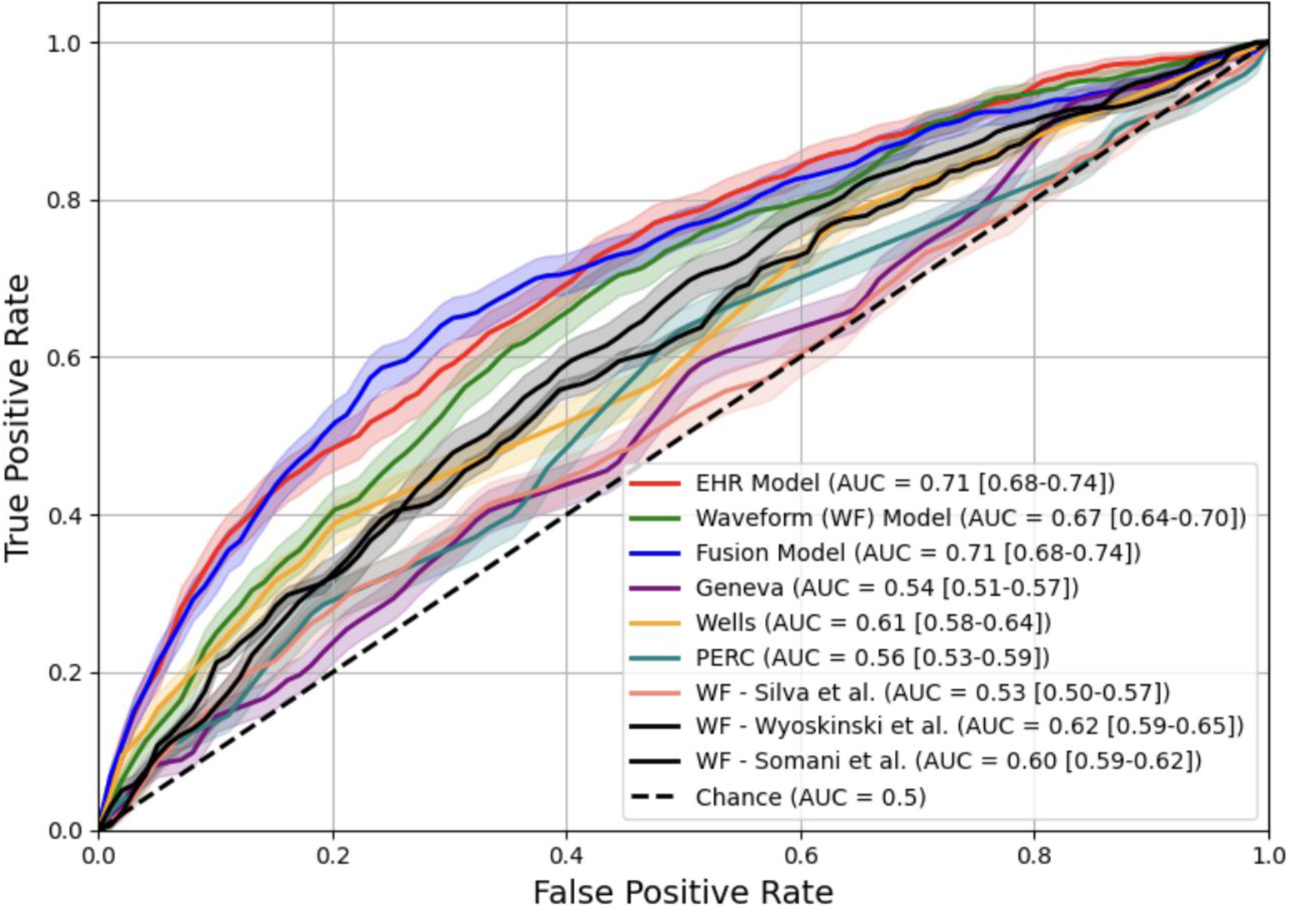
Test dataset AUROC values for all models trained in this study. The best performing stacked classifier model was selected as the Fusion model.

**Figure 3.**
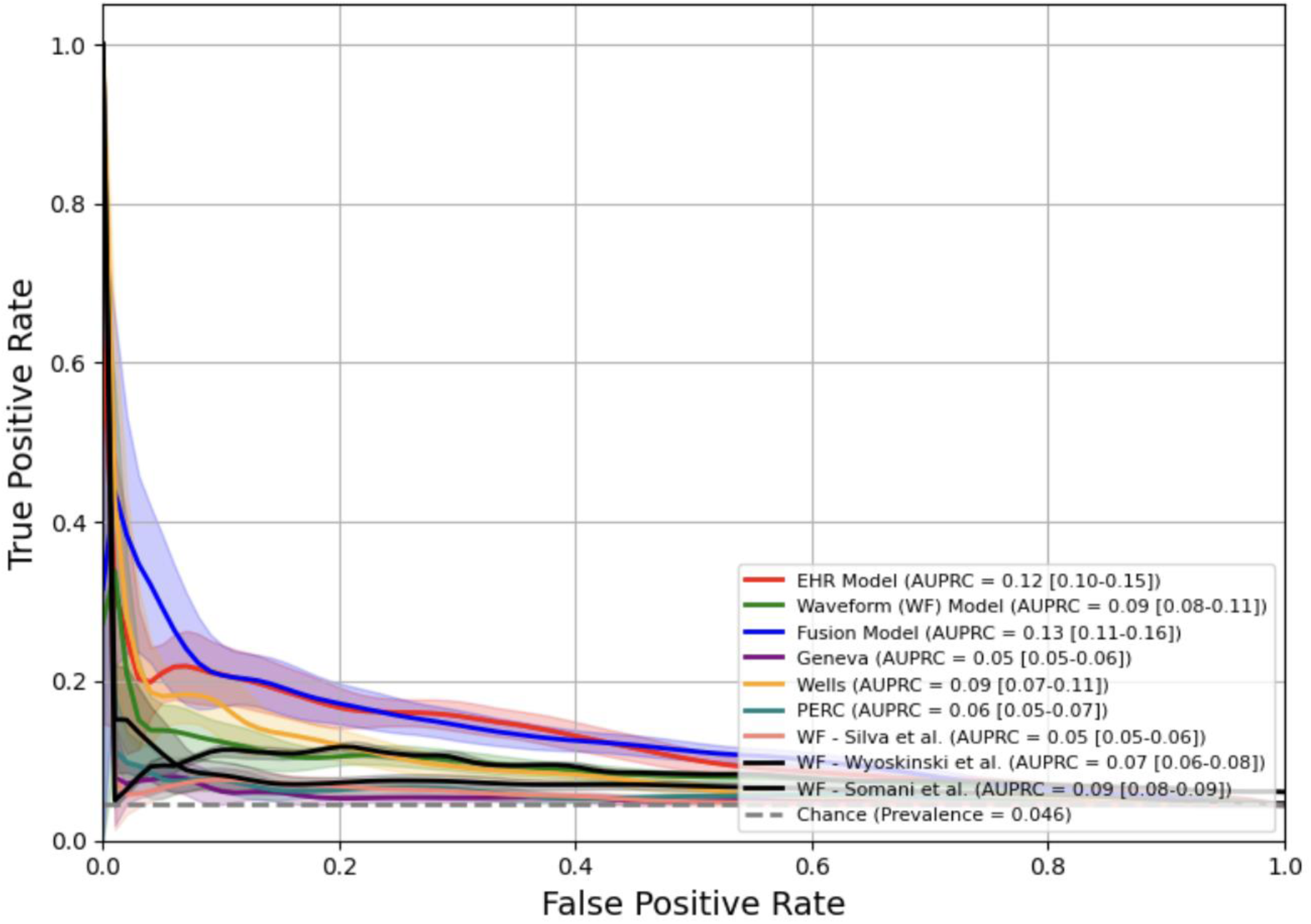
Test dataset AUPRC values for all models trained in this study. The best performing stacked classifier model was selected as the Fusion model. Results are presented with confidence intervals.

**Table 2:** Model performance metrics within the development dataset.

| Algorithm | AUROC (95% CI) | AUPRC (95% CI) | Brier Score (95% CI) | NPV** (95% CI) | PPV** (95% CI) | Specificity** (95% CI) |
| --- | --- | --- | --- | --- | --- | --- |
| **EHR Model** | **0.709 (0.679 – 0.738)** | 0.124 (0.101 – 0.152) | **0.042 (0.038 – 0.047)** | **0.980 (0.976 – 0.984)** | 0.068 (0.058 – 0.081) | **0.471 (0.400 – 0.541)** |
| **WF Model** | 0.668 (0.639 – 0.698) | 0.093 (0.075 – 0.115) | 0.043 (0.039 – 0.048) | 0.977 (0.972 – 0.982) | 0.061 (0.052 – 0.073) | 0.402 (0.336 – 0.490) |
| **Fusion Classifier** | 0.708 (0.677 – 0.739) | **0.132 (0.105 – 0.163)** | 0.063 (0.057 – 0.068) | 0.972 (0.966 – 0.977) | 0.055 (0.048 – 0.062) | 0.330 (0.280 – 0.382) |
| **Original Wells Score** | 0.607 (0.577 – 0.637) | 0.088 (0.070 – 0.110) | 0.044 (0.040 – 0.048) | 0.971 (0.965 – 0.978) | 0.049 (0.041 – 0.062) | 0.074 (0.000 – 0.369) |
| **Revised Geneva Score** | 0.542 (0.512 – 0.572) | 0.053 (0.045 – 0.063) | 0.065 (0.061 – 0.069) | 0.970 (0.960 – 0.981) | 0.050 (0.044 – 0.056) | 0.209 (0.170 – 0.276) |
| **Pulmonary Embolism Rule Out Criteria (PERC)** | 0.558 (0.526 – 0.590) | 0.058 (0.048 – 0.069) | 0.053 (0.049 – 0.057) | 0.956 (0.944 – 0.967) | 0.047 (0.041 – 0.052) | 0.172 (0.146 – 0.182) |
| **Wysokinski et al.** | 0.590 (0.561 – 0.620) | 0.069 (0.058 – 0.083) | 0.044 (0.039 – 0.048) | 0.973 (0.967 – 0.977) | 0.055 (0.048 – 0.062) | 0.338 (0.291 – 0.386) |
| **Somani et al.** | 0.603 (0.590 – 0.618) | 0.085 (0.079 – 0.091) | 0.264 (0.264 – 0.264) | 0.960 (0.956 – 0.963) | **0.069 (0.065 – 0.073)** | 0.303 (0.281 – 0.331) |
| **Silva et al.** | 0.559 (0.524 – 0.595) | 0.054 (0.046 – 0.064) | 0.058 (0.055 – 0.061) | 0.954 (0.946 – 0.964) | 0.046 (0.041 – 0.051) | 0.198 (0.171 – 0.247) |

\*note that the base classifiers for all models remain the same
\*\*thresholded metrics are calculated using a fixed sensitivity of 0.80 determined by clinician input.

### Feature Importance Analysis

Shapley values were aggregated across cross-validation folds to estimate feature importance under varying data splits. Figure 4 presents the waveform features in descending order of importance. Our analysis reveals several ECG features to be highly discriminative of PE, particularly those derived from vectorcardiography (VCG), heart rate variability (HRV), and Fourier analysis of the ECG waveform. Some of these biomarkers extend from known abnormalities, such as low-amplitude QRS complexes, while others like those from VCG remain relatively unexplored in the context of PE (40). While vectorcardiography has largely fallen out of clinical practice due to the ubiquity of the 12-lead ECG, it remains a valuable representation by capturing both the magnitude and direction of the heart’s electrical activity. This approach more effectively characterizes the heart’s spatiotemporal dynamics and can be leveraged for effective disease prediction (41). Our waveform classifier identified many HRV features as highly discriminative of PE. This observation comports with previous literature reporting a significant association between time-domain HRV metrics (i.e., SDNN) and right ventricular overload, a common finding in patients with PE (42). Other prototypical findings shared with the established literature include increased heart rate and alterations in the T-wave (43,44).

**Figure 4.**
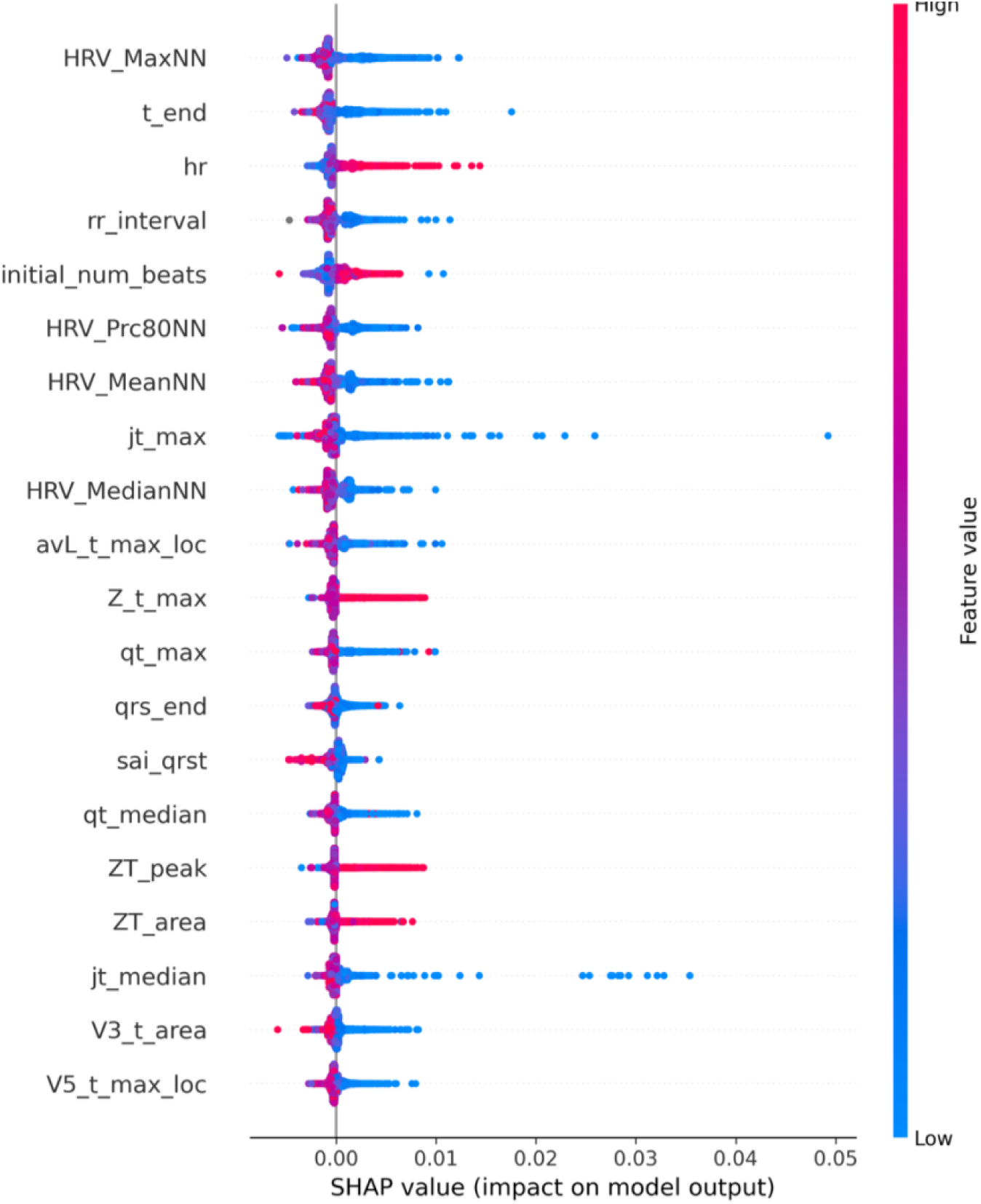
SHAP plot of the WF model trained in this study on data collected 48 hours prior to the time of PE suspicion.

Similarly, our EHR model feature importances are depicted in Figure 5, in decreasing order of feature importance. These features are generally concordant with previously described risk factors and symptomatic manifestations of PE, such as negative fluid balance and lowered blood pressure (i.e., hypotension) (45). Additionally, other factors we identify like increased mean heart rate and decreased oxygen saturation have long been recognized in the context of PE (46). Our SHAP analysis also identifies risk factors such as patient pain levels, Glasgow coma score (GCS), and Braden scale as being highly correlated with the occurrence of PE. These variables likely represent proxies for established risk factors. For example, low GCS often reflects sedation or neurologic injury with resultant immobility, which are well-recognized risk factors of PE (47).

**Figure 5.**
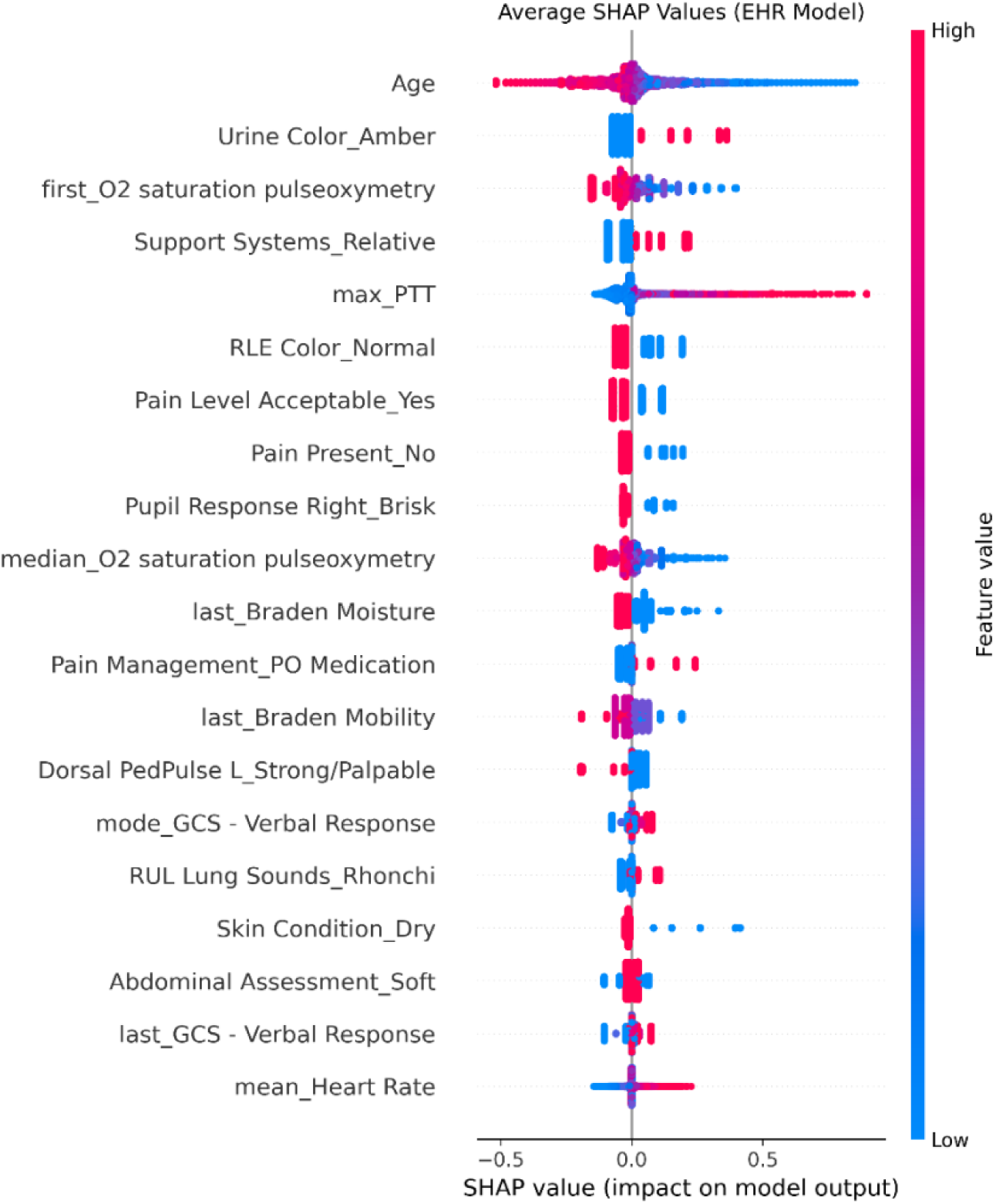
SHAP plot of the EHR model trained in this study on data collected 48 hours prior to the time of PE suspicion.

## Discussion

### Main findings

Using a large, open-source clinical dataset, we compared a WF, EHR-based, and fusion model for PE identification in patients with a moderate- to high suspicion of PE. To our knowledge, this study presents the first ECG waveform model for PE identification in an ICU-specific population. Our WF model is trained on 12-lead, 10-second ECG waveforms acquired at least 48 hours prior to a chest CT order and the EHR model is trained on clinical data from the same time window. We integrate these base classifier PE prediction probabilities from both modalities to create a stacked classifier which serves as our fusion model. Importantly, we design all models to be accessible in a variety of settings, incorporating only variables routinely collected during ICU admission. Notably, our WF model significantly outperforms several previously developed models and offers enhanced interpretability through a carefully curated and expansive waveform feature space. Similarly, our EHR and fusion models demonstrate superior overall performance to our selected benchmark clinical risk scores.

### Analysis of existing literature

The ECG waveform encodes a wealth of information about the heart’s physiological state, capturing key information of the spatiotemporal dynamics of electrical conduction across the atria and ventricles. These features are closely tied to a patient’s overall health and clinical outcomes, making ECG waveforms an information-rich modality for risk assessment and diagnosis. Machine learning (ML) models that leverage ECG waveforms can identify subtle patterns and correlations that may be overlooked by traditional methods, enhancing their diagnostic utility (48). These challenges emphasize the critical role of AI-based ECG diagnostics for PE in improving access to timely, effective care (49,50).

Several other studies have recognized this fact and utilized ECG waveform signals for the detection of PE in combination with other clinical data (18–20,39,51). However, many of these studies were designed with narrow predictive windows or specialized laboratory tests, limiting their actionability. In this study, we aimed to compare multiple approaches to identifying PE using only routine clinical data—a more challenging task but one with greater potential utility in proactive clinical decision-making. Following this aim, we intentionally exclude specialized laboratory tests (e.g., D-dimer) and utilize a longer lead time relative to previous approaches, collecting ECGs from 48 hours before a chest CT is ordered. This approach contrasts with prior studies, such as Somani et al. (20), where D-dimer was used to significantly enhance the performance of a fusion model combining laboratory, vital, and ECG data. Similarly, Wysokinski et al. (19) limited their ECG analysis to a narrow window of +/- 6 hours around a CTPA, restricting the actionable time frame for physicians. Su et al. (18) report high predictive performance with their ECG waveform model, but their study is limited by a small sample size and an unrealistic PE prevalence which limits generalizability. Other studies share similar constraints and were developed and validated on closed or proprietary datasets, limiting reproducibility. A key emphasis of our study is on interpretability of our models. To this end, we conduct a detailed explainability analysis of our handcrafted feature set, derived from both ECG and EHR data, using an open-source development dataset.

### Strengths

Deep neural networks (DNNs), including convolutional neural networks and, more recently, transformer-based models, have achieved considerable success in predictive tasks utilizing ECG waveform data. (52,53). However, a significant limitation of training DNNs lies in their dependence on large, diverse datasets to effectively generalize across different domains which poses a challenge when working with limited datasets such as ours. To address these limitations, we extracted over 1,000 expert-informed features from our ECG waveform data, thereby enhancing model interpretability, allowing us to reduce model complexity and outperform DNN models trained on our dataset. We also enhance clinical applicability of our models relative to previous studies by expanding the lead time of our model (i.e., limiting data to that acquired at more 48 hours prior to a chest CT order) and excluding specialized laboratory tests (e.g., D-dimer).

### Limitations

While all models demonstrated strong discrimination compared to our benchmark risk stratification models, the fusion model did not significantly outperform either the WF or EHR model contrary to our initial hypothesis. Furthermore, the fusion model has a markedly higher Brier score compared to either base model, indicating that the predicted probabilities are less reliable and limiting the model’s clinical utility. Another limitation of this study regards the uncertainty regarding the exact time of onset of PE, as the literature reports a mean diagnostic delay of over eight days (54). As a result, the actual onset may highly vary with a PE possibly occurring several days before our specified prediction window. To help address, this we attempted to utilize a broader predictive window than considered in earlier studies, utilizing ECGs waveforms up to 48 hours before the chest CT scan order to identify PE.

We also introduced sampling biases by utilizing a chest CT scan as a requirement to determine the time of PE clinical suspicion, ignoring patients who were categorized as low risk. However, we note that our study focuses on identification, which encompasses both detection and prediction tasks, and so we emphasize a long lead time to allow for more time for proactive treatment strategies or interventions. Because of this sampling process, we may also exclude some PE patients who may never receive a chest CT as they are contraindicated in a subset of patients (e.g., for those with renal disease) or may not be readily available due to resource constraints (55). This constraint may artificially reduce the PE sample to those with a high pretest probability, ignoring other patients with lower pretest probabilities and reducing the study sample size. Consequently, our model’s may not generalize to a broad, unselected ICU population without significant further validation. This limitation reframes the model’s potential clinical utility as a tool to improve risk stratification among PE patients already under the suspicion of PE rather than a tool for general screening. We also highlight the inability of traditional clinical risk scores to discriminate those with or likely to develop a PE within the ICU as these models were not designed for a critically ill population. Through this study, our WF model, using only a single 10-second ECG, achieves statistically significant performance improvements compared to baseline clinical risk scores like the Revised Geneva score and the PE Rule Out Criteria, while performing comparably to the Original Wells score.

Our lack of external validation may also raise questions regarding the generalizability of our models. We developed our approach using the largest publicly available clinical dataset that includes 500 Hz, 10-second ECG waveform data combined with corresponding EHR data; however, to our knowledge, no other open-source datasets offer both modalities, which limits opportunities for external validation. Second, as several other studies have noted, relying on ICD diagnostic codes to identify our cohort introduces potential inaccuracies—ICD-9 codes, for example, have a reported positive predictive value of only 29.9% for pulmonary embolism (56). Despite these challenges, our model performs robustly, and its feature importances align well with established risk factors in the literature. Future work will explore a refined label space, addressing the issues of ICD diagnostic coding and potentially improving cohort accuracy. Additional future directions include a prospective validation of our models at an independent site to validate their generalizability in real-world settings and further establish their clinical utility.

## Data Availability

All data produced in the present study are available upon reasonable request to the authors

## Author contributions statement

All authors conceived the experiments, S.R. conducted the experiments, all authors analyzed the results and reviewed the manuscript.

## Acknowledgements

This work was carried out at the Advanced Research Computing at Hopkins (ARCH) core facility (rockfish.jhu.edu), which is supported by the National Science Foundation (NSF) grant number OAC1920103.

This material is also based upon work supported by the National Science Foundation Graduate Research Fellowship under Grant No. DGE2139757, awarded to CH. Any opinion, findings, and conclusions or recommendations expressed in this material are those of the authors(s) and do not necessarily reflect the views of the National Science Foundation.

