## Supplement for "A Comparative Analysis of Electronic Health Record and Electrocardiogram Waveform Data for Pulmonary Embolism Identification in Critically Ill Patients"

**Table S1.** ICD 9 and 10 diagnostic codes used to define PE.

| ICD 9 Codes | 415.12, 415.12, 415.13, 415.19 |
| --- | --- |
| ICD 10 Codes | I26.0, I26.01, I26.02, I26.09, I26.90, I26.92, I26.93, I26.94, I26.99 |

### Table S2: Completed (Strengthening the reporting of observational studies in epidemiology) STROBE checklist

### Item

|  | **No** | **Recommendation** | **Completed C** |
| --- | --- | --- | --- |
| **Title and abstract** | 1 | (*a*) Indicate the study’s design with a commonly used term in the title or the abstract | **X** |
|  |  | (*b*) Provide in the abstract an informative and balanced summary of what was done and what was found | **X** |
| **Introduction** |  |  |  |
| Background/rationale | 2 | Explain the scientific background and rationale for the investigation being reported | **X** |
| Objectives | 3 | State specific objectives, including any prespecified hypotheses | **X** |
| **Methods** |  |  |  |
| Study design | 4 | Present key elements of study design early in the paper | **X** |
| Setting | 5 | Describe the setting, locations, and relevant dates, including periods of recruitment, exposure, follow-up, and data collection | **X** |
| Participants | 6 | (*a*) Give the eligibility criteria, and the sources and methods of selection of  participants. Describe methods of follow-up | **X** |
|  |  | (*b*) For matched studies, give matching criteria and number of exposed and unexposed | **N/A** |
| Variables | 7 | Clearly define all outcomes, exposures, predictors, potential confounders, and effect  modifiers. Give diagnostic criteria, if applicable | **X** |
| Data sources/ measurement | 8* | For each variable of interest, give sources of data and details of methods of assessment (measurement). Describe comparability of assessment methods if there is more than one group | **X** |
| Bias | 9 | Describe any efforts to address potential sources of bias | **X** |
| Study size | 10 | Explain how the study size was arrived at | **X** |
| Quantitative variables | 11 | Explain how quantitative variables were handled in the analyses. If applicable, describe which groupings were chosen and why | **X** |
| Statistical methods | 12 | (*a*) Describe all statistical methods, including those used to control for confounding | **X** |
|  |  | (*b*) Describe any methods used to examine subgroups and interactions | **X** |
|  |  | (*c*) Explain how missing data were addressed | **X** |
|  |  | (*d*) If applicable, explain how loss to follow-up was addressed | **X** |
|  |  | (*e*) Describe any sensitivity analyses | **X** |
| **Results** |  |  |  |
| Participants | 13* | (a) Report numbers of individuals at each stage of study—eg numbers potentially eligible, examined for eligibility, confirmed eligible, included in the study, completing follow-up, and analysed | **X** |
|  |  | (b) Give reasons for non-participation at each stage | **X** |
|  |  | (c) Consider use of a flow diagram | **X** |
| Descriptive data | 14* | (a) Give characteristics of study participants (eg demographic, clinical, social) and information on exposures and potential confounders | **X** |
|  |  | (b) Indicate number of participants with missing data for each variable of interest | **X** |
|  |  | (c) Summarise follow-up time (eg, average and total amount) | **N/A** |
| Outcome data | 15* | Report numbers of outcome events or summary measures over time | **X** |
| Main results | 16 | (*a*) Give unadjusted estimates and, if applicable, confounder-adjusted estimates and their precision (eg, 95% confidence interval). Make clear which confounders were adjusted for and why they were included | **X** |
|  |  | (*b*) Report category boundaries when continuous variables were categorized | **X** |
|  |  | (*c*) If relevant, consider translating estimates of relative risk into absolute risk for a  meaningful time period | **X** |

| Other analyses | 17 | Report other analyses done—eg analyses of subgroups and interactions, and  sensitivity analyses | **X** |
| --- | --- | --- | --- |
| **Discussion** |  |  |  |
| Key results | 18 | Present key elements of study design early in the paper | **X** |
| Limitations | 19 | Discuss limitations of the study, taking into account sources of potential bias or  imprecision. Discuss both direction and magnitude of any potential bias | **X** |
| Interpretation | 20 | Give a cautious overall interpretation of results considering objectives, limitations,  multiplicity of analyses, results from similar studies, and other relevant evidence | **X** |
| Generalisability | 21 | Discuss the generalisability (external validity) of the study results | **X** |
| **Other information** |  |  |  |
| Funding | 22 | Give the source of funding and the role of the funders for the present study and, if  applicable, for the original study on which the present article is based | **X** |

#### **Table S3:** Completed Transparent Reporting of a Multivariable Prediction Model for Individual Prognosis (TRIPOD) checklist

| Section/Topic | Item | Checklist item | Completed? |
| --- | --- | --- | --- |
| Title | 1 | Identify the study as developing and/or validating a multivariable prediction model, the target population, and the outcome to be predicted. | Yes |
| Abstract | 2 | Provide a summary of objectives, study design, setting, participants, sample size, predictors, outcome, statistical analysis, results, and conclusions. | Yes |
| Background | 3a | Explain the medical context (including whether diagnostic or prognostic) and rationale for developing or validating the multivariable prediction model, including references to  existing models. | Yes |
| Objectives | 3b | Specify the objectives, including whether the study describes the development or validation of the model, or both. | Yes |
| Source of Data | 4a | Describe the study design or source of data (e.g., randomized trial, cohort, or registry data), separately for the development and validation data sets, if applicable. | Yes |
|  | 4b | Specify the key study dates, including start of accrual; end of accrual; and, if applicable, end of follow-up. | Yes |
| Participants | 5a | Specify key elements of the study setting (e.g., primary care, secondary care, general population) including number and location of centres. | Yes |
|  | 5b | Describe eligibility criteria for participants. | Yes |
|  | 5c | Give details of treatments received, if relevant. | N/A because retrospective study. |
| Outcome | 6a | Clearly define the outcome that is predicted by the prediction model, including how and when assessed. | Yes |
|  | 6b | Report any actions to blind assessment of the outcome to be predicted. | N/A because retrospective study. |
| Predictors | 7a | Clearly define all predictors used in developing the multivariable prediction model, including how and when they were measured. | Yes |
|  | 7b | Report any actions to blind assessment of predictors for the outcome and other predictors. | Yes |
| Sample Size | 8 | Explain how the study size was arrived at. | Yes |
| Missing Data | 9 | Describe how missing data were handled (e.g., complete-case analysis, single imputation, multiple imputation) with details of any imputation method. | Yes |
| Statistical Analysis Methods | 10a | Describe how predictors were handled in the analyses. | Yes |
|  | 10b | Specify type of model, all model-building procedures (including any predictor selection), and method for internal validation. | Yes |
|  | 10c | For validation, describe how the predictions were calculated. | Yes |
|  | 10d | Specify all measures used to assess model performance and, if relevant, to compare multiple models. | Yes |
|  | 10e | Describe any model updating (e.g., recalibration) arising from the validation, if done. | N/A because no model updating. |
| Risk Groups | 11 | Provide details on how risk groups were created, if done. | N/A, not done. |
| Development vs. validation | 12 | For validation, identify any differences from the development data in setting, eligibility criteria, outcome, and predictors. | Yes |
| Participants | 13a | Describe the flow of participants through the study, including the number of participants with and without the outcome and, if applicable, a summary of the follow-up time. A diagram may  be helpful. | Yes |
|  | 13b | Describe the characteristics of the participants (basic demographics, clinical features, available predictors), including the number of participants with missing data for predictors and outcome. | Yes |
|  | 13c | For validation, show a comparison with the development data of the distribution of important variables (demographics, predictors, and outcome). | Yes |
| Model Development | 14a | Specify the number of participants and outcome events in each analysis. | Yes |
|  | 14b | If done, report the unadjusted association between each candidate predictor and outcome. | Yes |
| Model Specification | 15a | Present the full prediction model to allow predictions for individuals (i.e., all regression coefficients, and model intercept or baseline survival at a given time point). | Yes |
|  | 15b | Explain how to use the prediction model. | Yes |
| Model Performance | 16 | Report performance measures (with CIs) for the prediction model. | Yes |
| Model Updating | 17 | If done, report the results from any model updating (i.e., model specification, model performance). | Yes |
| Limitations | 18 | Discuss any limitations of the study (such as nonrepresentative sample, few events per predictor, missing data). | Yes |
| Interpretation | 19a | For validation, discuss the results with reference to performance in the development data, and any other validation data. | Yes |
|  | 19b | Give an overall interpretation of the results, considering objectives, limitations, results from similar studies, and other relevant evidence. | Yes |
| Implications | 20 | Discuss the potential clinical use of the model and implications for future research. | Yes |
| Supplementary Information | 21 | Provide information about the availability of supplementary resources, such as study protocol, Web calculator, and data sets. | Yes |
| Funding | 22 | Give the source of funding and the role of the funders for the present study. | Yes |

**Table S3.** All EHR features extracted and examined in the study

| Waveform Features* | 1. All features derived from electrocardiogram (ECG), heart rate variability, and ECG-derived respiration analyses using NeuroKit2 (1) 2. All features derived from ECG-derived vectorcardiogram analysis using Braveheart (2) 3. All features derived from lead II data of the 12-lead ECG using general time series analysis implemented with tsfresh (3) |
| --- | --- |

*Due to the high number of features across different types of analyses, we refer to each analytical package that was used to create each type of feature. Documentation for each package describes all available features that can be extracted.

**Table S4.** All EHR features extracted and examined in the study

| Category or Variable | Specific Processing and Features Extracted |
| --- | --- |
| Demographics |  |
| Age | Ages greater than 90 were converted to 90 due to dataset limitations |
| Sex | Male or female, one-hot encoded |
| Ethnicity | One-hot encoded |
| Admission Information |  |
| Admission Source | One-hot encoded |
| ICU Type | One-hot encoded |
| Height | On admission |
| Weight | First value after ICU admission |
| Physiological Scoring |  |
| Glasgow Coma Score (total, verbal, motor, eyes) | Mode, Min, Max First, Last recorded value |
| RASS | Mode, Min, Max First, Last recorded value |
| Braden Scale (sensory perception, moisture, activity, mobility, nutrition, friction and shear) | Mode, Min, Max First, Last recorded value |
| Medical History |  |
| Dialysis | Binary flag |
| Stroke | Binary flag |
| Hypertension | Binary flag |
| Deep Vein Thrombosis | Binary flag |
| Pulmonary Embolism | Binary flag |
| Congestive Heart Failure | Binary flag |
| Myocardial Infarction | Binary flag |
| Hypercoagulability Disorder | Binary flag |
| Heritable Clotting Disorder | Binary flag |
| Arrhythmia | Binary flag |
| Chronic Respiratory Disorder | Binary flag |
| Osteoarthritis | Binary flag |
| Paralysis | Binary flag |
| Cancer | Binary flag |
| Diabetes | Binary flag |
| Transplant | Binary flag |
| Transfusion | Binary flag |
| Cirrhosis | Binary flag |
| Hemoptysis | Binary flag |
| Fracture | Binary flag |
| Labs |  |
| Albumin | Mean, minimum, maximum, first, last, median, slope |
| Alkaline Phosphate | Mean, minimum, maximum, first, last, median, slope |
| Ammonia | Mean, minimum, maximum, first, last, median, slope |
| Amylase | Mean, minimum, maximum, first, last, median, slope |
| Anion Gap | Mean, minimum, maximum, first, last, median, slope |
| AST/ALT Ratio | Mean, minimum, maximum, first, last, median, slope |
| Band Neutrophils | Mean, minimum, maximum, first, last, median, slope |
| Base Deficit | Mean, minimum, maximum, first, last, median, slope |
| Base Excess | Mean, minimum, maximum, first, last, median, slope |
| Basophil (%) | Mean, minimum, maximum, first, last, median, slope |
| Bedside Glucose | Mean, minimum, maximum, first, last, median, slope |
| Bicarbonate | Mean, minimum, maximum, first, last, median, slope |
| Bilirubin (Total, Direct, Indirect) | Mean, minimum, maximum, first, last, median, slope |
| Blood Urea Nitrogen | Mean, minimum, maximum, first, last, median, slope |
| BUN/Creatinine Ratio | Mean, minimum, maximum, first, last, median, slope |
| Calcium | Mean, minimum, maximum, first, last, median, slope |
| Carboxyhemoglobin | Mean, minimum, maximum, first, last, median, slope |
| Chloride | Mean, minimum, maximum, first, last, median, slope |
| Creatinine | Mean, minimum, maximum, first, last, median, slope |
| Eosinophil (%) | Mean, minimum, maximum, first, last, median, slope |
| Fibrinogen | Mean, minimum, maximum, first, last, median, slope |
| Glucose | Mean, minimum, maximum, first, last, median, slope |
| Hematocrit | Mean, minimum, maximum, first, last, median, slope |
| Hemoglobin | Mean, minimum, maximum, first, last, median, slope |
| High-density Lipoprotein | Mean, minimum, maximum, first, last, median, slope |
| Ionized Calcium | Mean, minimum, maximum, first, last, median, slope |
| Iron | Mean, minimum, maximum, first, last, median, slope |
| Lactate | Mean, minimum, maximum, first, last, median, slope |
| Low-density Lipoprotein | Mean, minimum, maximum, first, last, median, slope |
| Lymphocytes (%) | Mean, minimum, maximum, first, last, median, slope |
| Mean Corpuscular Hemoglobin | Mean, minimum, maximum, first, last, median, slope |
| Mean Corpuscular Hemoglobin Concentration | Mean, minimum, maximum, first, last, median, slope |
| Mean corpuscular volume | Mean, minimum, maximum, first, last, median, slope |
| Mean platelet volume | Mean, minimum, maximum, first, last, median, slope |
| Methemoglobin | Mean, minimum, maximum, first, last, median, slope |
| Monocytes (%) | Mean, minimum, maximum, first, last, median, slope |
| O2 Saturation | Mean, minimum, maximum, first, last, median, slope |
| Oxyhemoglobin | Mean, minimum, maximum, first, last, median, slope |
| paCO2 | Mean, minimum, maximum, first, last, median, slope |
| paO2 | Mean, minimum, maximum, first, last, median, slope |
| Partial Thromboplastin Time | Mean, minimum, maximum, first, last, median, slope |
| pH | Mean, minimum, maximum, first, last, median, slope |
| Phosphate | Mean, minimum, maximum, first, last, median, slope |
| Platelets | Mean, minimum, maximum, first, last, median, slope |
| Polymorphonuclear Leukocytes (%) | Mean, minimum, maximum, first, last, median, slope |
| Potassium | Mean, minimum, maximum, first, last, median, slope |
| Protein C | Mean, minimum, maximum, first, last, median, slope |
| Protein S | Mean, minimum, maximum, first, last, median, slope |
| Prothrombin Time | Mean, minimum, maximum, first, last, median, slope |
| Prothrombin Time - International Normalized Ratio | Mean, minimum, maximum, first, last, median, slope |
| Red Blood Cells | Mean, minimum, maximum, first, last, median, slope |
| Red Cell Distribution Width | Mean, minimum, maximum, first, last, median, slope |
| Serum Alanine Transaminase | Mean, minimum, maximum, first, last, median, slope |
| Serum Aspartate Transaminase | Mean, minimum, maximum, first, last, median, slope |
| Serum Osmolality | Mean, minimum, maximum, first, last, median, slope |
| Sodium | Mean, minimum, maximum, first, last, median, slope |
| Thyroid Stimulating Hormone | Mean, minimum, maximum, first, last, median, slope |
| Thyroxine | Mean, minimum, maximum, first, last, median, slope |
| Total Cholesterol | Mean, minimum, maximum, first, last, median, slope |
| Total Protein | Mean, minimum, maximum, first, last, median, slope |
| Transferrin | Mean, minimum, maximum, first, last, median, slope |
| Triglycerides | Mean, minimum, maximum, first, last, median, slope |
| Troponin - I | Mean, minimum, maximum, first, last, median, slope |
| Troponin - T | Mean, minimum, maximum, first, last, median, slope |
| Urinary Creatinine | Mean, minimum, maximum, first, last, median, slope |
| Urinary Sodium | Mean, minimum, maximum, first, last, median, slope |
| Urinary Specific Gravity | Mean, minimum, maximum, first, last, median, slope |
| White Blood Cells in Urine | Mean, minimum, maximum, first, last, median, slope |
| White Blood Cells | Mean, minimum, maximum, first, last, median, slope |
| Medications |  |
| Acetaminophen | Binary indicator of administration |
| Adrenergic Bronchodilators | Binary indicator of administration |
| Aminoglycosides | Binary indicator of administration |
| Anticholinergic Bronchodilators | Binary indicator of administration |
| Anticholinergics | Binary indicator of administration |
| Antidiarrheals | Binary indicator of administration |
| Antiemetics | Binary indicator of administration |
| Antihistamines | Binary indicator of administration |
| Barbiturates | Binary indicator of administration |
| Benzodiazepines | Binary indicator of administration |
| Beta Blockers | Binary indicator of administration |
| Calcium Channel Blockers | Binary indicator of administration |
| Carbapenems | Binary indicator of administration |
| Cephalosporins | Binary indicator of administration |
| Class V Antiarrhythmics | Binary indicator of administration |
| Colloid fluids | Binary indicator of administration |
| Crystalloid fluids | Binary indicator of administration |
| Diuretics | Binary indicator of administration |
| General Anesthetics | Binary indicator of administration |
| Glucocorticoids | Binary indicator of administration |
| Glucose Elevating Drugs | Binary indicator of administration |
| Glycopeptides | Binary indicator of administration |
| H2 Receptor Blockers | Binary indicator of administration |
| Insulin | Binary indicator of administration |
| Laxatives | Binary indicator of administration |
| Lincomycins | Binary indicator of administration |
| Macrolides | Binary indicator of administration |
| Monoamine oxidase inhibitors Antidepressants | Binary indicator of administration |
| Methylxanthines | Binary indicator of administration |
| Neuromuscular Blockers | Binary indicator of administration |
| NSAIDs | Binary indicator of administration |
| Opioids | Binary indicator of administration |
| Other antidepressants | Binary indicator of administration |
| Beta-Lactams | Binary indicator of administration |
| Phenylpiperazine Antidepressants | Binary indicator of administration |
| Potassium Channel Blockers | Binary indicator of administration |
| Proton Pump Inhibitor | Binary indicator of administration |
| Quinolones | Binary indicator of administration |
| Serotonin and Norepinephrine Reuptake Inhibitor Antidepressants | Binary indicator of administration |
| Sodium Channel Blockers | Binary indicator of administration |
| Somatostatin | Binary indicator of administration |
| SSRI Antidepressants | Binary indicator of administration |
| Sulfonamides | Binary indicator of administration |
| Tetracyclic Antidepressants | Binary indicator of administration |
| Tetracyclines | Binary indicator of administration |
| Tricyclic Antidepressants | Binary indicator of administration |
| Vasodilators | Binary indicator of administration |
| Vasopressors | Binary indicator of administration |
| Physiological Measurements |  |
| Temperature | Mean, minimum, maximum, first, last, median, slope |
| Blood Pressure | tsfresh package features |
| SaO2 | tsfresh package features |
| Respiratory Rate | tsfresh package features |
| Heart Rate | tsfresh package features |
| Urine Output | Volume in mL over last 24 hours |
| Urine Color | One-hot encoded |
| Pupil Response | One-hot encoded |
| Dorsal Pedal Pulse | One-hot encoded |
| Abdominal Assessment | One-hot encoded |
| Skin Condition | One-hot encoded |
| Treatments |  |
| Cryoprecipitate transfusion | Binary indicator |
| Packed red blood cell transfusion | Binary indicator |
| Plasma transfusion | Binary indicator |
| Platelet transfusion | Binary indicator |
| Surgery | Binary indicator |
| Comorbidities |  |
| AIDS/HIV | Binary indicator from Elixhauser ICD codes |
| Alcohol Abuse | Binary indicator from Elixhauser ICD codes |
| Blood Loss Anemia | Binary indicator from Elixhauser ICD codes |
| Cardiac Arrhythmias | Binary indicator from Elixhauser ICD codes |
| Chronic Pulmonary Disease | Binary indicator from Elixhauser ICD codes |
| Coagulopathy | Binary indicator from Elixhauser ICD codes |
| Congestive Heart Failure | Binary indicator from Elixhauser ICD codes |
| Deficiency Anemia | Binary indicator from Elixhauser ICD codes |
| Depression | Binary indicator from Elixhauser ICD codes |
| Diabetes, Complicated | Binary indicator from Elixhauser ICD codes |
| Diabetes, Uncomplicated | Binary indicator from Elixhauser ICD codes |
| Drug Abuse | Binary indicator from Elixhauser ICD codes |
| Fluid and Electrolyte Disorders | Binary indicator from Elixhauser ICD codes |
| Hypertension, Complicated | Binary indicator from Elixhauser ICD codes |
| Hypertension, Uncomplicated | Binary indicator from Elixhauser ICD codes |
| Hypothyroidism | Binary indicator from Elixhauser ICD codes |
| Liver Disease | Binary indicator from Elixhauser ICD codes |
| Lymphoma | Binary indicator from Elixhauser ICD codes |
| Metabolic Acidosis | Binary indicator from lab data |
| Metastatic Cancer | Binary indicator from Elixhauser ICD codes |
| Obesity | Binary indicator from Elixhauser ICD codes |
| Other Neurological Disorders | Binary indicator from Elixhauser ICD codes |
| Paralysis | Binary indicator from Elixhauser ICD codes |
| Peptic Ulcer Disease | Binary indicator from Elixhauser ICD codes |
| Peripheral Vascular Disorders | Binary indicator from Elixhauser ICD codes |
| Renal Failure | Binary indicator from Elixhauser ICD codes |
| Rheumatoid Arthritis/Collagen Vascular Diseases | Binary indicator from Elixhauser ICD codes |
| Solid Tumor Without Metastasis | Binary indicator from Elixhauser ICD codes |
| Valvular Disease | Binary indicator from Elixhauser ICD codes |
| Weight Loss | Binary indicator from Elixhauser ICD codes |
| Other |  |
| Support Systems | One-hot encoded |
| RLE Color | One-hot encoded |
| Pain Level Acceptable | Binary flag |
| Pain Present | Binary flag |
| Pain Management | One-hot encoded |
| RUL Lung Sounds | One-hot encoded |

**Table S5.** Stacked Classifier Model Performances Within the Fusion Model

| Stacked Classifier Algorithm in Fusion Model* | AUROC (95% CI) | AUPRC (95% CI) | Brier Score (95% CI) | NPV** (95% CI) | PPV** (95% CI) | Specificity** (95% CI) |
| --- | --- | --- | --- | --- | --- | --- |
| XGBoost | 0.650 (0.619 – 0.681) | 0.090 (0.073 – 0.110) | 0.060 (0.055 – 0.065) | 0.968 (0.958 – 0.977) | 0.052 (0.045 – 0.061) | 0.280 (0.215 – 0.385) |
| Random Forest | 0.690 (0.660 – 0.720) | 0.115 (0.092 – 0.143) | 0.050 (0.045 – 0.054) | **0.981 (0.977 – 0.984)** | **0.070 (0.060 – 0.084)** | **0.484 (0.427 – 0.571)** |
| Logistic Regression | 0.664 (0.635 – 0.694) | 0.107 (0.083 – 0.135) | 0.045 (0.041 – 0.049) | 0.976 (0.970 – 0.980) | 0.059 (0.051 – 0.068) | 0.384 (0.309 – 0.445) |
| CatBoost | **0.708 (0.677 – 0.739)** | **0.132 (0.105 – 0.163)** | **0.063 (0.057 – 0.068)** | 0.972 (0.966 – 0.977) | 0.055 (0.048 – 0.062) | 0.330 (0.280 – 0.382) |

*all models have the same baseline classifier predictions as inputs

**sensitivity fixed at 0.80 for the thresholded metrics

**Table S6.** EHR Model Performances across all tested models

| EHR Model Type* | AUROC (95% CI) | AUPRC (95% CI) | Brier Score (95% CI) | NPV** (95% CI) | PPV** (95% CI) | Specificity** (95% CI) |
| --- | --- | --- | --- | --- | --- | --- |
| XGBoost | 0.677 (0.645 – 0.708) | **0.130 (0.102 – 0.164)** | **0.044 (0.039 – 0.049)** | 0.977 (0.972 – 0.980) | 0.061 (0.053 – 0.069) | 0.401 (0.347 – 0.456) |
| Random Forest | 0.696 (0.666 – 0.726) | 0.102 (0.084 – 0.121) | 0.043 (0.039 – 0.047) | 0.979 (0.974 – 0.983) | 0.065 (0.056 – 0.076) | 0.439 (0.370 – 0.511) |
| Logistic Regression | **0.709 (0.679 – 0.738)** | 0.124 (0.101 – 0.152) | 0.042 (0.038 – 0.047) | **0.980 (0.976 – 0.984)** | **0.068 (0.058 – 0.081)** | **0.471 (0.400 – 0.541)** |
| CatBoost | 0.661 (0.628 – 0.693) | 0.104 (0.083 – 0.128) | **0.044 (0.040 – 0.048)** | 0.974 (0.967 – 0.979) | 0.058 (0.049 – 0.067) | 0.365 (0.284 – 0.436) |

**sensitivity fixed at 0.80 for the thresholded metrics

**Table S7.** WF Model Performances across all tested models

| WF Model Type* | AUROC (95% CI) | AUPRC (95% CI) | Brier Score (95% CI) | NPV** (95% CI) | PPV** (95% CI) | Specificity** (95% CI) |
| --- | --- | --- | --- | --- | --- | --- |
| XGBoost | 0.614 (0.582 - 0.646) | 0.088 (0.068 - 0.111) | **0.045 (0.040 – 0.050)** | 0.970 (0.962 – 0.977) | 0.054 (0.046 – 0.063) | 0.316 (0.246 – 0.392) |
| Random Forest | **0.668 (0.639 – 0.698)** | **0.093 (0.075 – 0.115)** | 0.043 (0.039 – 0.048) | **0.977 (0.972 – 0.982)** | **0.061 (0.052 – 0.073)** | **0.402 (0.336 – 0.490)** |
| Logistic Regression | 0.589 (0.559 - 0.619) | 0.061 (0.051 - 0.071) | **0.045 (0.040 – 0.049)** | 0.971 (0.964 – 0.977) | 0.054 (0.047 – 0.062) | 0.322 (0.258 – 0.381) |
| CatBoost | 0.620 (0.588 - 0.651) | 0.079 (0.065 - 0.095) | 0.044 (0.040 – 0.049) | 0.970 (0.964 – 0.976) | 0.053 (0.047 – 0.060) | 0.309 (0.263 – 0.367) |

**sensitivity fixed at 0.80 for the thresholded metrics

**Table S6:** Modifications made to Geneva reference model features

| **Original Feature** | **Modified Feature** | **Reason** |
| --- | --- | --- |
| Age > 65 y | No modification | N/A |
| Previous DVT or PE | No modification | N/A |
| Surgery (under general anesthesia) or fracture (of the lower limbs within 1 mo | Any surgical procedure or fracture of the lower limbs | Lack of documentation clarifying general anesthesia |
| Active malignant condition (solid or hematologic malignant condition, currently active or considered cured < 1 y) | Any record of malignancy within the medical record in addition to currently active diagnosis | Lack of documentation of whether malignancy was considered cured |
| Unilateral lower-limb pain | N/A | Not available reliably within the dataset |
| Hemoptysis | No modification | N/A |
| Heart Rate: 75–94 beats/min | No modification | N/A |
| Heart Rate: >= 95 beats/min | No modification | N/A |
| Pain on lower-limb deep venous palpation and unilateral edema | N/A | Not available reliably within the dataset |

**Table S7:** Modifications made to Wells reference model features

| **Original Feature** | **Modified Feature** | **Reason** |
| --- | --- | --- |
| Clinical signs & symptoms DVT | Recorded diagnosis of DVT | Lack of documentation of all clinical signs and symptoms of DVT |
| Tachycardia (> 100/min) | No modification | N/A |
| Immobilization or surgery in the previous four weeks | Any surgical procedure or fracture of the lower limbs | More precisely defined |
| Previous DVT/PE | No modification | N/A |
| Hemoptysis | No modification | N/A |
| Malignancy | No modification | N/A |
| An alternative diagnosis is less likely than PE | N/A | Information not reliably available within the dataset |

**Table S9.** Comparison of current model to prior PE waveform prediction models

| Prediction Model | Years of Study | Primary Outcome | PE development dataset prevalence | Development dataset  Sample Size | ECG Collection Time | Patient Description | Internal AUROC | Hospitals in Study |
| --- | --- | --- | --- | --- | --- | --- | --- | --- |
| Current Model | 2008-2019 | Prediction of PE in critically ill inpatients | 4.60% | 66,239 | 48 hours prior to imaging study order | Age >= 18, in ICU, had a Chest CT scan | 0.67 | 1 |
| SPPH-ECG (4) | 2018-2021 | Prediction of PE in hospitalized patients | 49.70% | 658 | 48 hours from the onset of patient symptoms | Age >= 18, had a CTPA, in ICU, no serious primary pulmonary disease and associated pulmonary hypertension | 0.87 | 1 |
| Wysokinski et al. (5) | 1999-2020 | Prediction of PE in hospitalized patients | 9.3% | 79,894 | The closest ECG to the CTPA date within ±6 hours | Age >= 18, had a CTPA report and ECG | 0.69 | 16 |
| Silva et al. (6) | 2017-2021 | Prediction of PE in emergency department patients | 37%^a^ | 1,014 | Not stated | Age >= 18, had a CTPA,  No SARS-COV2 infection diagnosis, D-dimer acquired  within 12 hours before CTPA | 0.75 | 1 |
| Somani et al. (7) | 2003-2020 | Prediction of PE in hospitalized patients | 20.2% | 21, 183 | PE-positive ECGs acquired within 24 hours of CTPA time. PE-negative ECGs acquired prior to 6 months of a PE-positive CTPA or anytime from a PE-negative CPTA study. | Age >= 18, had a CTPA, no neurological disease, no sedation | 0.59 | 5 |

^a^Only Validation data prevalence is provided
